# Personalizing neuromodulation for chronic pain: A connectivity-guided randomized trial

**DOI:** 10.64898/2026.03.02.26347430

**Authors:** Enrico De Martino, Margit Midtgaard Bach, Bruno Nascimento Couto, Anne Jakobsen, Pedro Nascimento Martins, Stian Ingemann-Molden, Adenauer G. Casali, Thomas Graven-Nielsen, Daniel Ciampi de Andrade

**Affiliations:** Center for Neuroplasticity and Pain (CNAP), Department of Health Science and Technology, Faculty of Medicine, Aalborg University, Aalborg, Denmark; Federal University of Juiz de Fora, Juiz de Fora, Brazil; Institute of Science and Technology, Federal University of São Paulo, São Paulo, Brazil

**Keywords:** Non-invasive cortical stimulation, neuromodulation, cortical connectivity, biomarkers of treatment response, precision medicine, neuropathic pain, nociplastic pain, nociceptive pain

## Abstract

In this randomized, double-blind, controlled trial of 8 weeks of repetitive transcranial magnetic stimulation (rTMS) for chronic pain, we compared the classic primary motor cortex (M1) rTMS with a novel target-selection strategy based on pre-therapy cortical connectivity. Guided by principles of homeostatic plasticity, we tested whether stimulating the cortical site with the lowest pre-therapy global connectivity would be more effective than two active comparators: stimulating the site with the highest pre-therapy global connectivity or stimulating M1 independent of connectivity. Before starting rTMS treatment, TMS-evoked EEG potentials were recorded from four cortical targets: M1, the dorsolateral prefrontal cortex, the anterior cingulate cortex, and the posterosuperior insular cortex. For each target, global connectivity was quantified using a distance-weighted, phase-based index (debiased weighted phase lag index, wPLI) derived from pre- and post-TMS-evoked EEG activity, capturing both the magnitude and spatial extent of TMS-induced oscillatory phase locking across cortical regions. Target allocation in the Low- and High-Connectivity groups was based on this global connectivity measure. Ninety patients with chronic pain were randomized to Low-Connectivity, High-Connectivity, or Classic-M1 groups. Treatment consisted of 12 rTMS sessions delivered over 8 weeks to the assigned target. The primary outcome was the proportion of patients achieving ≥ 30% reduction in pain intensity. Secondary outcomes included continuous change in pain intensity, pain interference, sleep, fatigue, mood, quality of life, and patient global impression of change. No between-group differences were observed for primary or secondary outcomes (p > 0.05). In exploratory analyses, we examined whether pre-therapy local connectivity (within-target wPLI) predicted treatment response. In the Classic-M1 group, lower pre-therapy local M1 connectivity was associated with a greater reduction in pain intensity (r = 0.50, p = 0.005). This association was not observed in the Low- or High-Connectivity groups. A regression model including group-by-connectivity interaction indicated that the relationship between local connectivity and pain reduction differed between the Classic-M1 and High-Connectivity groups (p = 0.038). The results of this clinical trial showed that connectivity-based target allocation using global connectivity did not improve clinical outcomes. However, lower local M1 connectivity was associated with greater pain reduction following Classic-M1 stimulation, suggesting that local M1 connectivity may serve as a potential biomarker of response.

## Introduction

Chronic pain is one of the most prevalent and disabling medical conditions^1^, and its therapeutic management is challenging, as most treatments show modest to moderate effects in clinical trials^2^. In the absence of methods to individualize treatments and offer interventions to those more likely to respond, a ‘trial-and-error’ approach remains the predominant strategy in clinical practice, often leading to delays in effective pain control^3^. These limitations indicate the need for a more individualized and precision-medicine approach to pain management.^4^

Repetitive transcranial magnetic stimulation (rTMS) has emerged as a promising intervention for alleviating chronic pain.^5^ TMS uses brief, high-intensity magnetic pulses to induce electric currents, thereby stimulating neurons by depolarizing myelinated axons in the cortex,^6^ with low side effects. rTMS targeting the primary motor cortex (M1) is the most extensively studied approach in clinical trials,^7^ due to its effects on pain-modulatory networks.^8^ Several studies have shown analgesic effects of rTMS M1 in peripheral neuropathic pain and fibromyalgia, with a response rate between 35% and 50% of patients.^9–12^ A smaller number of studies have reported analgesic effects of rTMS targeting three non-motor cortical regions: the left dorsolateral prefrontal cortex (DLPFC), anterior cingulate cortex (ACC), and posterosuperior insula (PSI). The DLPFC is a key region for mood and cognitive-appraisal aspects of pain.^13^ rTMS of the DLPFC has been used with some success for the treatment of chronic pain associated with co-morbid depressive symptoms,^14^ migraine,^15^ burning mouth syndrome,^16^ and peripheral neuropathic pain.^7^ ACC is a key region of the limbic system and is involved in cognitive and affective aspects of pain processing.^17,18^ rTMS has shown that targeting the ACC can alleviate chronic pain in fibromyalgia,^19^ while also leading to a reduction in negative mood and anxiety scores.^20^ PSI is a major cortical recipient of spinothalamic projections and plays a key role in processing incoming sensory-discriminative nociceptive signals and modulating nociceptive gain.^21^ rTMS of the PSI produced analgesia in patients with chronic peripheral neuropathic pain.^22^ Despite these findings, response rates to rTMS across all four stimulation targets generally remain below 50%, with responders experiencing an average pain reduction of approximately 30%.^9–12^ This modest efficacy may reflect substantial inter-individual variability in pain processing, as cortical areas can be differentially engaged and modulated by pain experience across individuals.^23^

In this context, we propose that cortical connectivity measures derived from TMS can provide mechanistic insight into the inter-individual variability in rTMS response. By combining TMS with electroencephalography (TMS-EEG), we found that phase-based connectivity metrics derived from TMS-evoked potentials (TEPs) may be relevant for treatment optimization. Specifically, we showed that reduced phase-based connectivity measures, including inter-trial phase coherence (ITC)^24^ and weighted Phase Lag Index (wPLI),^25^ were associated with individual pain sensitivity,^26^ and that rTMS targeting M1 increased both measures, with effects persisting beyond the stimulation session.^27^

These findings led us to investigate cortical phase-based connectivity as a potential neurophysiological predictor of rTMS response. In the feasibility pilot study of this clinical trial, we found that pre-therapy cortical connectivity, quantified using a distance-weighted debiased wPLI index, showed the potential to predict the pain-reducing response to M1 rTMS^28^. People with chronic pain with lower pre-therapy global connectivity were more likely to respond to ten sessions of M1 rTMS with reduced pain, suggesting that reduced connectivity before treatment may indicate a greater capacity for rTMS to modulate cortical dynamics.^28^ Given the established pain-modulatory effects of M1 stimulation, enhanced M1 responsiveness to rTMS may translate into more effective pain reduction, supporting the potential use of pre-therapy TMS-EEG to guide rTMS target selection.

Building on this rationale, the present study aimed to determine whether rTMS efficacy could be improved by guiding target selection based on pre-therapy cortical connectivity. We conducted a trial using TMS-EEG to quantify connectivity among four candidate targets: M1, left DLPFC, ACC, and PSI. Patients were randomized to one of three groups. In the Low-Connectivity and High-Connectivity groups, the pre-therapy connectivity index was calculated for each target and used to identify, respectively, the cortical site with the lowest or highest global connectivity as the stimulation target. In the Classic-M1 group, rTMS was delivered to M1 independent of the pre-therapy connectivity measures. We hypothesized that the use of the cortical areas with the lowest pre-therapy global connectivity as a target for rTMS would lead to a higher number of responders compared to two active-control groups: one in which rTMS was delivered to the target with the highest connectivity, and a second in which therapy was delivered to the Classic-M1 target, independently of the results of pre-therapy global connectivity measures.^7,10,12^

## Materials and methods

### Participants

The study was conducted at the Center for Neuroplasticity and Pain, Aalborg University, Aalborg (Denmark) from April 2024 to June 2025. The study was approved by the local ethics committee “North Denmark Region Committee” (N-20230076), registered on ClinicalTrials.gov (NCT06395649), and the protocol was registered on the Open Science Framework website (https://osf.io/b2e8h/overview).^29^ Ninety patients with chronic pain participated in a double-blind, randomized, controlled, three-arm, parallel-group trial. Patients were eligible to participate if they were between 18 and 80 years old and had chronic pain, defined as pain present on most days for more than 3 months. Patients were required to report a mean pain intensity of 3-9 on a numeric rating scale (NRS; 0=no pain; 10=worst pain imaginable). Participants were excluded if they were pregnant or breastfeeding, had uncontrolled major depression as the primary diagnosis, or had current or recent history of substance abuse. Furthermore, exclusion criteria included an inability to cooperate with study procedures or complete study assessments due to cognitive difficulties, as well as any contraindication to TMS, such as a history of epilepsy, the presence of cranial ferromagnetic implants (e.g., intracranial neurostimulators), and tattoos or permanent makeup containing metallic pigments in the facial region. Patients were also excluded if they were unable to fulfill the criteria in the Transcranial Magnetic Stimulation Adult Safety Screen questionnaire.^30^ All potential patients underwent a standardized screening process prior to enrolment, during which pain mechanisms (neuropathic, nociceptive, or nociplastic)^31^ and ICD-11 pain categories (primary or secondary pain)^32^ were determined.

### Sample size estimation

The sample size calculation was informed by previous studies demonstrating analgesic effects of rTMS applied to the four cortical targets examined in this protocol. In a trial on neuropathic pain, Attal et al. reported that M1-rTMS had a 44% response rate and decreased pain intensity by 1.5 ± 0.8 numerical rating scale (NRS) points after 8 sessions, compared with 0.8 ± 1.5 NRS points under sham stimulation.^7^ Short et al. found that 10 sessions of left DLPFC rTMS in fibromyalgia patients yielded an average daily pain intensity reduction of around 29% relative to baseline, compared with around 4% in the sham group.^14^

In an rTMS study targeting the ACC in fibromyalgia, Tzabazis et al. observed 31–56% reductions in average pain intensity after 20 sessions of 10-Hz rTMS, compared with 15% reductions with sham stimulation.^19^ Dongyang et al. demonstrated that five sessions of 10-Hz rTMS to PSI in peripheral neuropathic pain resulted in a 58% response rate, with reduced visual analogue scale (VAS) scores from 68 to 34 mm (≈50% reduction) compared with 66 to 52 mm (≈24% reduction) under sham.^22^ Based on these findings, the analgesic effects of rTMS across the four cortical targets in chronic pain can be characterized as moderate (Cohen’s d = 0.4–0.5). The present study assumed a standardized mean difference (Cohen’s d) of 0.4 for the contrast between cortical regions with low and high connectivity, with a negligible therapeutic effect in the latter condition (i.e., sham stimulation). For the three-arm parallel design, a two-sided significance level of α = 0.05 was used, with a step-down Dunnett correction to control the family-wise error rate across the two experimental vs control comparisons. The disjunctive power was set at 1 − β = 0.80 under the global alternative hypothesis, with clinically relevant effect sizes defined as δ[= 0.65 (clinically meaningful improvement, corresponding to 30% reduction in pain intensity) and δ[= 0.12 (negligible improvement). Assuming equal allocation across all three arms, these assumptions yield a required sample size of n=79 patients, with a family-wise error rate of 0.05 and a disjunctive power of 0.80. Anticipating a dropout rate of approximately 15%, we therefore planned to recruit 90 patients, divided into three groups of 30 patients each.

### Procedure

The patients were allocated in a 1:1:1 ratio to one of three treatment arms: Low-Connectivity, High-Connectivity, or Classic-M1. Each participant was randomized to one of these three groups prior to inclusion using a computerized random sequence generator (https://www.randomizer.org), ensuring balanced allocation with a planned sample of 30 patients per arm. For participants assigned to the Low-Connectivity group, rTMS was applied to the individually identified cortical region with the lowest global connectivity. For those assigned to the High-Connectivity group, rTMS was applied to the cortical region with the highest global connectivity. Participants assigned to the Classic-M1 group received stimulation over M1 according to established protocols,^7^ without consideration of connectivity, although still assessed. Patients and assessors were blinded to group assignment throughout the trial. All patients underwent an assessment 1 week before starting rTMS treatment, including questionnaires and a connectivity assessment using TMS-EEG.

For the rTMS intervention, the induction phase consisted of five consecutive daily rTMS sessions delivered over one week. This was followed by a 7-week maintenance phase with one rTMS session per week (12 sessions in total). Each session lasted 20 minutes, including 15 minutes of active stimulation and 5 minutes of questionnaires. Target locations remained the same throughout the treatment. This protocol was similar to previous rTMS studies demonstrating analgesic effects in patients with chronic pain.^7,33^ Patients were allowed to keep their concomitant analgesic drug treatment throughout the study.

### Transcranial Magnetic Stimulation evoked electroencephalographic responses

Single-pulse TMS was applied in a randomized order to the four cortical regions, M1, DLPFC, ACC, and PSI (Figure 1), following previously established procedures.^23,27,28^ Stimulation was delivered using a biphasic stimulator (MagPro R30, MagVenture A/S, Denmark) equipped with a figure-of-eight coil (Cool-B65) and a double-cone coil (Cool-D-B80). TMS-evoked potentials were acquired with a TMS-compatible passive 63-channel EEG cap (g.tec medical engineering GmbH, Austria), with the ground electrode placed on the right zygoma, Cz at the vertex, and the reference on the right mastoid^27,34^. The electrode impedances were kept below 5 kΩ, and the data were sampled at 4.800 Hz (g.HIamp amplifier, g.tec medical engineering GmbH).^35^ To minimize auditory evoked potentials from the TMS coil click, patients wore noise-cancelling in-ear headphones (ER3C, Etymotic Research, Schaumburg, USA) in combination with the TMS Auditory Artifact Control (TAAC) masking toolbox.^36^ To minimize somatosensory contamination from coil vibration, two net caps (GVB-geliMED GmbH, Germany) and a thin plastic stretch wrap were placed over the EEG cap.^23,27^ Coil positioning was guided using neuronavigation (InVesalius Navigator, version 3.1), with the participant’s head and the TMS coil calibrated through an optical infrared tracking system (Polaris Vicra, NDI, Ontario, Canada).^37^ The navigated TMS system continuously monitored coil location and maintained targeting accuracy below 3 mm.^38^ The rt-TEP toolbox supported real-time visualization and quality control of TEPs.^39^ For M1, PSI, and ACC stimulation was delivered over the hemisphere contralateral to the side of greatest pain intensity. If no side predominance was reported, the left hemisphere was selected. The M1 hotspot for the hand was determined by identifying the coil position that elicited the largest visible hand twitch, typically corresponding to C3 (left) or C4 (right). The rest motor threshold (rMT) was defined as the lowest intensity producing a visible hand twitch.^40^ The left DLPFC target was located in the middle frontal gyrus using the method described previously.^41^ This location corresponds to the area below the F3 electrode, and an intensity of 110% of the hand rMT was used.^42^ For the ACC, the target was localized 4 cm anterior to the tibialis anterior (TA) muscle motor hotspot,^43^ corresponding to Fc1 (left) or Fc2 (right). The TA muscle hotspot was identified as the coil position eliciting a visible leg muscle twitch. The TA rMT was defined as the lowest intensity required to elicit this response. ACC TMS-EEG was performed at 90% of the TA rMT. The PSI target was determined using a validated fast-PSI localization formula.^44^ The electrodes C5 or C6 were nearest to the stimulation site. PSI stimulation was delivered at 90% of the TA rMT. To ensure clear TEPs, defined as a peak-to-peak amplitude between 6 µV and 25 µV within the 20–100 ms window, the coil orientation and stimulation intensity were adjusted for each cortical target, and the averaged TEPs were calculated across the first 30 stimuli. This process was used to evoke a clear local cortical response while minimizing excessive cortical spread.

**Figure 1:**
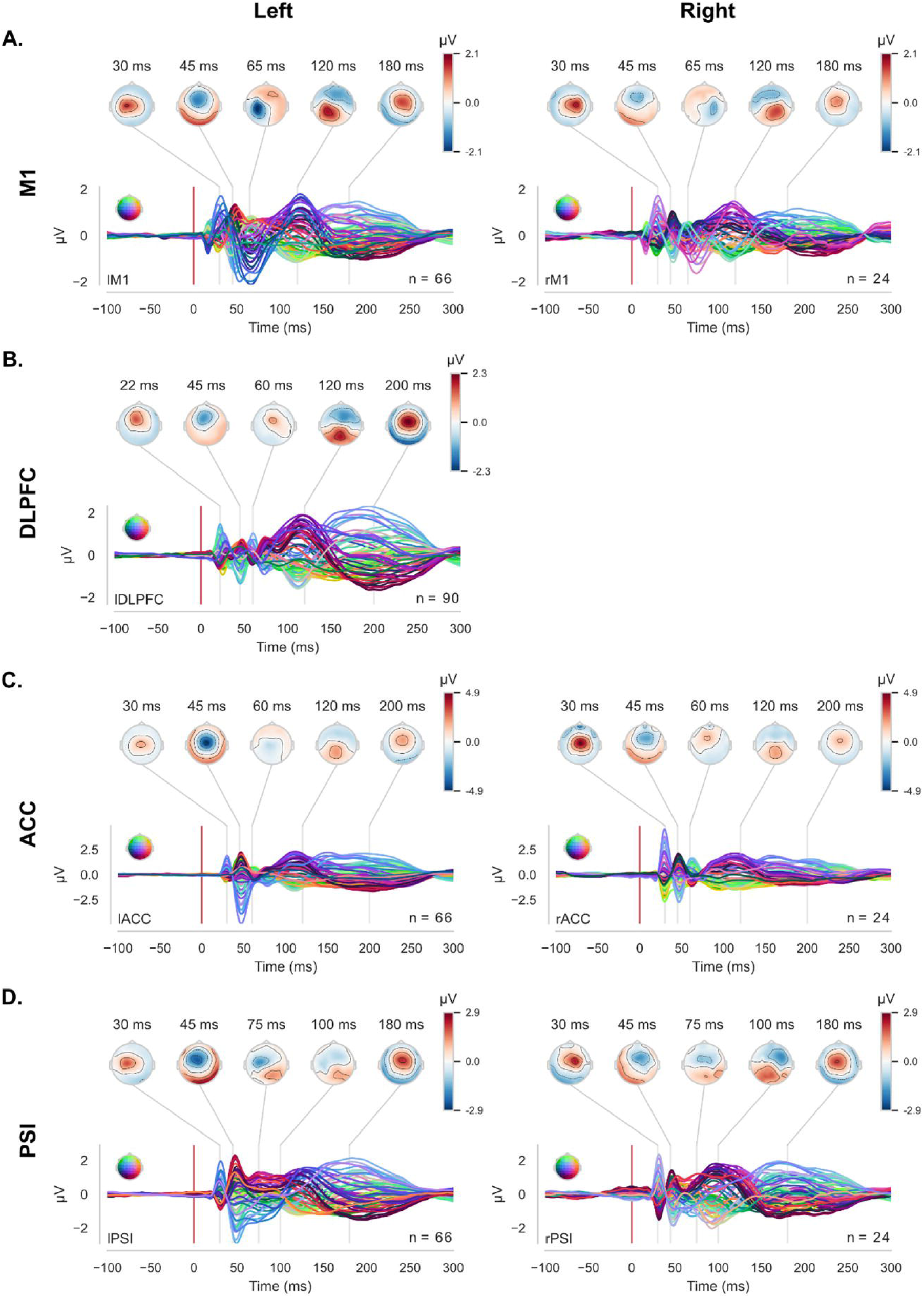
Grand-average TMS-evoked EEG potentials (TEPs). **(A)** primary motor cortex (M1), **(B)** dorsolateral prefrontal cortex (DLPFC), **(C)** anterior cingulate cortex (ACC), and **(D)** posterior superior insula (PSI) for left- and right-hemisphere. For each cortical target, the lower panels display butterfly plots of TEPs from all EEG channels averaged across patients. The red vertical line indicates the TMS pulse at 0 ms. Above each butterfly plot, topographical maps illustrate the spatial distribution of TEP amplitudes at the characteristic post-stimulation time.

EEG preprocessing was performed using a custom-designed Python pipeline (Python Software Foundation). Continuous EEG data were segmented into epochs from −800 to +800 ms relative to the TMS trigger.^28^ To correct for the TMS-induced artifact, the signal segment from 0 to 15 ms (for M1 and DLPFC stimulation) and from 0 to 25 ms (for ACC and PSI stimulation) was replaced with a mirrored copy of the pre-stimulus data.^27^ Epochs and channels contaminated by decay artifacts, 50 Hz line noise, ocular activity, or muscle activity were visually inspected and manually removed.^28^ Independent component analysis (fast-ICA) was subsequently applied to eliminate any remaining artifacts.^45,46^ The data were then band-pass filtered between 1 and 45 Hz using a third-order Butterworth filter, downsampled to 1200 Hz, and re-referenced to the average reference^28^, and cleaned by interpolating removed channels using spherical spline interpolation.^47^

#### Classification of the targets using the connectivity index

The global connectivity index used to classify targets was extracted from the pre- and post-TMS stimulus periods for all electrode pairs using the debiased wPLI.^48^ EEG data were downsampled to 300 Hz, and time–frequency decomposition was performed using Morlet wavelets with 3.5 cycles, spanning 15 logarithmically spaced frequencies from 8 to 35 Hz. Pre-TMS stimulus connectivity was estimated within a window from −500 to −50 ms relative to the TMS pulse, whereas post-TMS connectivity was assessed between 30 and 130 ms. Connectivity changes were first computed as the normalized mean difference between connectivity at pre- and post-TMS periods. To quantify the spatial propagation of TMS-induced changes in connectivity, a distance-weighted wPLI propagation index was computed. For each electrode pair, the peak TMS-induced change in connectivity was identified as the 95^th^ percentile of the post–pre difference. These values were then weighted by the Euclidean distance between electrode pairs, and the median of the weighted peaks across all pairs was calculated. This global connectivity index was developed in a pilot study^28^ as a standardized measure integrating both the magnitude and spatial extent of TMS-induced connectivity, thereby reducing the likelihood that the metric would be disproportionately driven by local response amplitude and allowing direct comparisons across distinct stimulation targets.

To assess whether local connectivity values across targets were primarily driven by differences in local cortical excitability, local mean field power (LMFP) was also computed within stimulation-specific EEG clusters (Supplementary Table 1), and a Spearman correlation analysis was performed to assess whether the connectivity index correlated with LMFP. LMFP was defined as the root-mean-square of TEPs across predefined electrode clusters surrounding the stimulation site,^49^ within the same time window used to compute the connectivity index.

### Treatment by repetitive transcranial magnetic stimulation

The therapeutic intervention consists of 10-Hz rTMS applied to one of the four targets. Target locations were determined during the TMS–EEG session and recorded under neuronavigation using three anatomical distances from external landmarks (distance from the vertex, distance from the tragus, and diagonal distance).^50^ The rTMS treatment consisted of thirty trains, each lasting 10 seconds, with 20-second inter-train intervals, and delivered in accordance with standard procedures. Each train contained 100 pulses, for a total of 3000 administered over 15 minutes. Stimulation intensity varied according to the cortical target: 90% of the rMT of the hand muscle for M1,^7^ 110% of the rMT of the hand muscle for DLPFC,^51^ and 90% of the leg muscle PSI^22^ and ACC.^19^ These protocols have been used in previous studies involving patients with chronic pain and have demonstrated efficacy and safety.^7,14,19,22^

### Blinding assessment

At the end of the last treatment session, blinding efficacy was evaluated using a structured set of questions administered to patients and rTMS operators. The assessment aimed to determine whether blinding was maintained throughout the trial. Participants and operators were asked whether they believed they could identify the treatment sequence to which the participant had been allocated, and which treatment condition they believed had been administered. Previous studies have demonstrated that this type of structured inquiry provides a robust evaluation of blinding integrity.^52^

### Primary outcome

The primary outcome was the proportion of participants who reported a ≥30% reduction in VAS pain intensity at the last visit (week 8) compared with the average VAS pain intensity during the 7 days prior to study initiation. Pain intensity was measured using a VAS, where 0 represents “no pain” and 100 represents “the worst pain imaginable.”^53^

### Secondary outcome

Secondary outcomes included the proportion of participants who reported a ≥50% reduction in VAS pain intensity at the last visit compared with the average VAS pain intensity during the 7 days prior to study initiation. Furthermore, the following outcomes were assessed before and at the end of the study: pain intensity over the last 24 hours and the last 7 days (assessed by VAS), Brief Pain Inventory (BPI), sleep quality, fatigue, mood and anxiety symptoms (Hospital Anxiety and Depression Scale, HADS), health-related quality of life (EuroQol-5D, EQ-5D), and global impression of change assessed by both patients (PGIC) and operators (OGIC). The BPI was administered to quantify both pain severity and pain interference. Pain severity was calculated as the mean of four intensity items (worst, least, average, and current pain), and pain interference was calculated as the mean of seven items assessing the impact of pain on general activity, mood, walking ability, normal work, relationships with others, sleep, and enjoyment of life.^54^ The spatial distribution of pain was quantified using body maps,^55^ which were divided into 29 predefined anatomical regions. The total number of regions marked as painful was calculated to assess the extent of the pain.^56^ Mood and anxiety symptoms were assessed using the Hospital Anxiety and Depression Scale (HADS), a 14-item self-report measure covering anxiety (7 items) and depression (7 items), with higher scores indicating greater symptom severity.^57^ Health-related quality of life was evaluated using the EuroQol-5D (EQ-5D), which assesses five dimensions: mobility, self-care, usual activities, pain/discomfort, and anxiety/depression.^58^ Finally, sleep quality and fatigue were measured using two VAS. For sleep, participants rated “How did you sleep last night?” on a scale from 0 (bad sleep) to 100 (good sleep). For fatigue, they rated “How tired have you been in the last 24 hours?” from 0 (no tiredness) to 100 (worst imaginable tiredness).^59^ Patient Global Impression of Change (PGIC) and Operator Global Impression of Change (OGIC) were assessed at the end of the maintenance phase using standard 7-point Likert scales ranging from “very much worse” to “very much improved,” and were compared across treatment groups.

### Explorative analyses

Using the wPLI values to compute the connectivity index for patient classification, we additionally derived a measure of local connectivity at the target site. This was calculated within stimulation-specific EEG electrode clusters used for the LMFP as the average debiased wPLI across all electrode pairs within each cluster, thereby reflecting within-cluster phase-based connectivity.

### Statistical analyses

Descriptive statistics were used to characterize the distribution of all variables. Continuous variables were summarized using means and standard deviations (SD) or medians and interquartile ranges, as appropriate, based on their distribution (Shapiro–Wilk test), while categorical variables were summarized using absolute and relative frequencies. A two-sided significance level of α = 0.05 was used for all analyses.

A primary intention-to-treat analysis was conducted, with all randomized participants analyzed in their assigned treatment group. The primary outcome, responder status, was analyzed using a chi-squared test to compare proportions between treatment groups. In addition, an explorative intention-to-treat sensitivity analysis was conducted, including only participants who attended at least four induction sessions. Furthermore, to assess whether contrasts between the randomized stimulation target and M1 predicted treatment response, the connectivity index and local-to-M1 difference (LTM1D) variable were computed as the difference between target-area and M1 connectivity. Treatment response was analyzed using an ordinal logistic regression model with pain VAS reduction at the end of the trial as the outcome.

Secondary clinical outcomes were analyzed using baseline-adjusted regression models. Ordinal outcomes were analyzed using ordinal logistic regression, while continuous outcomes were analyzed using linear regression. All models included fixed effects for treatment group, time, and a treatment-by-time interaction, as well as a nonlinear term for baseline pain intensity to account for potential nonlinearity in baseline–outcome relationships. PGIC and OFIC were analyzed using the Kruskal–Wallis’ test.

Exploratory analyses examined associations between local connectivity values and changes in VAS pain intensity from baseline to the end of treatment, assessed over the previous 24 hours and the previous week. Linear regression models were fitted to assess the relationship between connectivity measures and pain reduction (percentage reduction from baseline), including interaction terms to evaluate whether these associations differed by stimulation target. This analysis was conducted in an exploratory, hypothesis-generating manner.

## Results

### Patient characteristics

A total of 132 patients were screened for eligibility, of whom 90 were included (Figure 2). Two patients had the baseline assessment but did not receive any rTMS treatment. Baseline demographic and clinical characteristics are summarized in Table 1. The three arms were broadly comparable in age, sex distribution, pain mechanism, ICD-11 pain classification, and years with pain. In the High-Connectivity arm, 57% of participants were assigned to PSI, followed by ACC (23%) and M1 (17%), with DLPFC selected in only one case (1%). In contrast, in the Low Connectivity arm, 53% of participants were assigned to DLPFC, followed by M1 (27%), with PSI and ACC each accounting for 10%. Most participants across groups received rTMS on the left hemisphere. Patient’s primary pain diagnoses are shown in Supplementary Table 2. Across all three groups, patients exhibited a broad range of chronic pain conditions, typical of clinical chronic pain populations. Diagnoses were described as either primary or secondary pain syndromes according to the ICD-11 classification and also included the mechanistic classification of pain: neuropathic conditions (e.g., radiculopathies, polyneuropathies, nerve lesions), nociceptive and degenerative musculoskeletal disorders (e.g., osteoarthritis, spondyloarthropathies, arthropathies), and nociplastic pain syndromes (e.g., fibromyalgia, chronic primary cervical pain, and chronic widespread pain). Medication used before and after rTMS treatment is summarized in Supplementary Table 3. The medication was similar across the three groups before and after the rTMS treatment, and no substantial between-group differences were observed. Across all groups, the most frequently used medications were acetaminophen, NSAIDs, pregabalin/gabapentin, and weak opioids.

**Figure 2:**
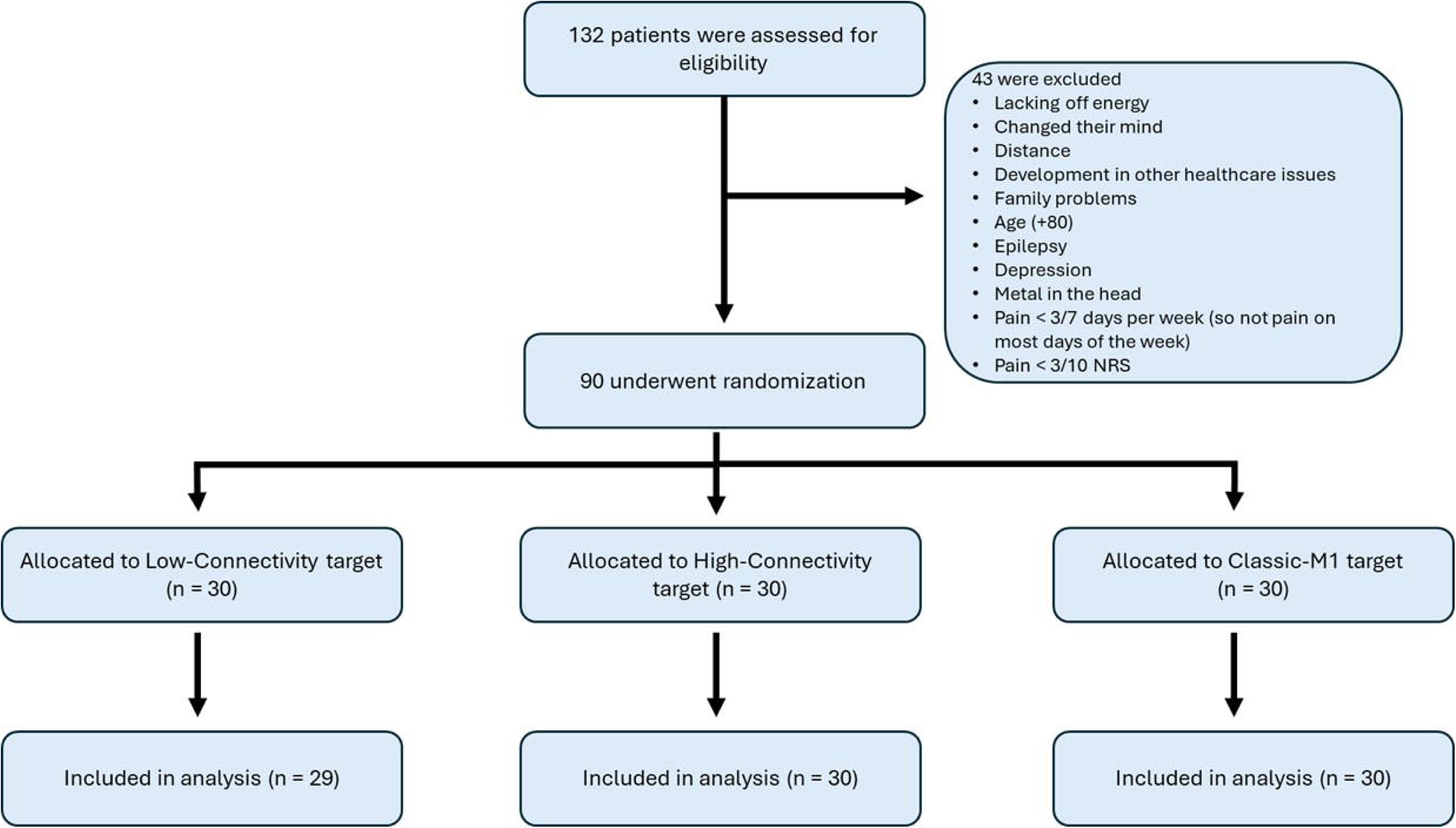
CONSORT flow diagram. 132 patients were assessed for eligibility; 43 were excluded, and 90 were randomized in a 1:1:1 ratio to the Low-Connectivity, High-Connectivity, or Classic-M1 arm. All participants allocated to the High-Connectivity and Classic-M1 groups were included in the final analysis, whereas one participant in the Low-Connectivity group was excluded.

**Table 1:** Baseline characteristics of the study groups.

| Characteristic |  | Classic-M1<br>N = 30 | High-Connectivity<br>N = 30 | Low-Connectivity<br>N = 30 |
| --- | --- | --- | --- | --- |
| Age (years)<sup>1</sup> |  | 51 (42-62) | 55 (46, 60) | 57 (48, 61) |
| Sex | Male | 12 (40%) | 11 (37%) | 9 (30%) |
|  | Female | 17 (57%) | 19 (63%) | 21 (70%) |
|  | Intersex | 1 (3.3%) | 0 (0%) | 0 (0%) |
| Mechanism-based<br>classification | Neuropathic | 10 (33%) | 9 (30%) | 10 (33%) |
|  | Nociceptive | 10 (33%) | 11 (37%) | 9 (30%) |
|  | Nociplastic | 10 (33%) | 10 (33%) | 11 (37%) |
| IDC-11<br>classification | Primary pain | 11 (37%) | 10 (33%) | 10 (33%) |
|  | Secondary pain | 19 (63%) | 20 (67%) | 20 (67%) |
| Years with pain |  | 9 (5, 15) | 7 (3, 15) | 9 (4, 16) |
| rTMS target | M1 | 30 (100%) | 5 (17%) | 8 (27%) |
|  | DLPFC | 0 (0%) | 1 (3%) | 16 (53%) |
|  | ACC | 0 (0%) | 7 (23%) | 3 (10%) |
|  | PSI | 0 (0%) | 17 (57%) | 3 (10%) |
| Hemisphere<br>contralateral to<br>most pain | Left | 24 (80%) | 22 (73%) | 20 (67%) |
|  | Right | 6 (20%) | 8 (27%) | 10 (33%) |

<sup>1</sup>Age is presented as median (interquartile range).
All other variables are presented as n (%), with percentages calculated within each group.

### Transcranial magnetic stimulation intensities, compliance, and side effects

TMS intensities used for TMS-EEG and rTMS are reported in Supplementary Table 4. Out of the 12 total rTMS sessions, the average number of rTMS sessions attended was 11.2±2.0 (96%) in the Low-Connectivity group, 11.4 ± 1.2 (96%) in the High-Connectivity group, and 11.8±0.3 (98%) in the Classic-M1 group. One patient in the High-Connectivity group missed six consecutive sessions (from weeks 3 to 8), and one in the Low-Connectivity group missed five consecutive sessions (from weeks 4 to 8). All other missed sessions were isolated single sessions or two consecutive missed sessions (7 in the High-Connectivity group, 9 in the Low-Connectivity group, and 6 in the Classic-M1 group). The most common reasons for missed sessions were seasonal diseases or personal reasons. Side effects reported during the rTMS treatment period are summarized in Supplementary Table 5. Overall, rTMS was well-tolerated across all three treatment arms, with no serious adverse events reported. The most common side effect was headache, occurring in 30% of participants in the Classic-M1 group, 40% in the High-Connectivity group, and 37% in the Low-Connectivity group. Dizziness was also relatively frequent, reported by 13% of participants in the Classic-M1 group, 27% in the High-Connectivity group, and 37% in the Low-Connectivity group. Other side effects, such as twitching, numbness, or electrical sensations, unusual sensory experiences, or feelings of confusion or drowsiness, were reported infrequently (≤13% in all groups), with no significant differences between treatment arms (all p>0.17). Reports of unusual smells, tastes, or emotions were uncommon, occurring in 0–10% of participants across groups. No significant side effects were observed between the treatment groups, and all adverse events were mild and transient. Most side effects (59%) were reported during the induction week, and 41% during the maintenance phase.

### Blinding assessment

Forty-two participants (38%) reported they could identify the treatment group to which they had been allocated. However, only 9 of these correctly identified their assigned group. Regarding therapist blinding, in 12 treatment cases (13%), the rTMS therapist reported being able to determine the patient’s group. However, only 4 of these guesses were correct.

### Primary outcome

Fourteen patients in the Low-Connectivity group (48%), 15 in the High-Connectivity group (50%), and 11 in the Classic-M1 group (37%) showed a ≥30% reduction in pain VAS scores at the end of treatment. The percentage reduction in pain was 33±55% in the Low-Connectivity group, 32±35% in the High-Connectivity group, and 27±33% in the Classic-M1 group (Figure 3). In the intention-to-treat analysis, there was no significant difference in responder rates between treatment groups for either the categorical or continuous analysis (Table 2). Results were unchanged in a modified intention-to-treat sensitivity analysis that included participants who attended at least 4 induction sessions (p = 0.475). No significant associations were observed between the global connectivity index and changes in VAS pain ratings, and no significant interactions with Time or treatment group were detected (Supplementary Table 6). Similarly, in the analysis examining the LTM1D difference, no significant main effects or interactions were found (Supplementary Table 6). Global connectivity values are reported in Supplementary Table 7.

**Figure 3:**
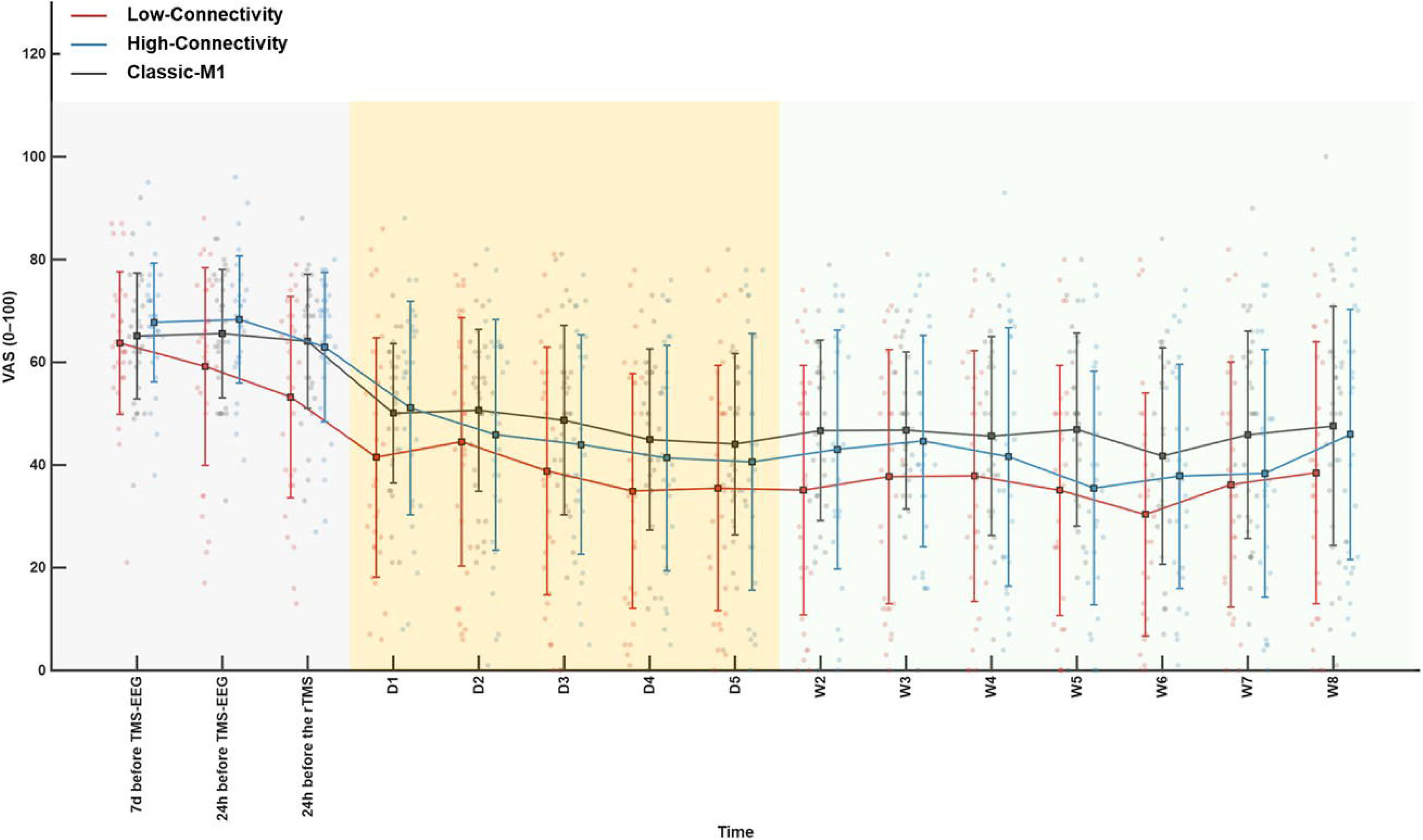
Pain intensity scores on a visual analogue scale. Data points represent individual participants, and square markers indicate group means with error bars representing standard deviations. Pain intensity VAS scores (0-100) were assessed at baseline time points (7 days before TMS–EEG, 24 hours before TMS–EEG, and 24 hours before rTMS), during the induction phase (D1–D5; yellow shaded area), and throughout the maintenance phase, with pain assessed at the time of each visit (W2–W8; green shaded area).

**Table 2:** Primary and secondary outcomes.

Primary and secondary outcomes
| Outcome | Before rTMS treatment |  |  | After rTMS treatment |  |  | p-value |
| --- | --- | --- | --- | --- | --- | --- | --- |
|  | Classic-M1 | High-Connectivity | Low-Connectivity | Classic M1 | High connectivity | Low connectivity |  |
| Primary outcome<br>(number of responders) | - | - | - | 11 (37%) | 15 (50%) | 14 (48%) | 0.530<sup>1</sup> |
| Secondary outcomes |  |  |  |  |  |  |  |
| Last 7 days (VAS) | 64 (59, 75) | 68 (65, 73) | 64 (57, 73) | 58 (40, 72) | 62 (40, 76) | 47 (35, 64) | 0.346 |
| Last 24 h (VAS) | 67 (57, 76) | 69 (61, 75) | 64 (52, 75) | 63 (34, 72) | 62 (35, 78) | 54 (37, 65) | 0.188 |
| BPI |  |  |  |  |  |  |  |
| Pain severity | 22 (18, 25) | 24 (21, 28) | 22 (18, 26) | 20 (14, 25) | 20 (17, 26) | 18 (14, 22) | 0.581 |
| Pain interference | 45 (34, 50) | 37 (27, 46) | 43 (32, 47) | 38 (28, 45) | 36 (18, 40) | 34 (27, 46) | 0.954 |
| Sleep VAS | 37 (26, 65) | 41 (24, 75) | 34 (21, 65) | 56 (37, 80) | 67 (33, 77) | 72 (40, 85) | 0.180 |
| Fatigue VAS | 70 (58, 76) | 66 (29, 74) | 66 (52, 77) | 60 (40, 66) | 58 (30, 70) | 60 (50, 69) | 0.724 |
| HADS | 14 (11, 18) | 11 (3, 15) | 11 (7, 15) | 12 (6, 15) | 9 (5, 15) | 9 (4, 15) | 0.286 |
| EQ-5D | 12 (11, 15) | 12 (10, 13) | 12 (10, 14) | 12 (10, 13) | 11 (10, 13) | 11 (9, 12) | 0.259 |
| PGIC | - | - | - | 3 (3, 4) | 3 (3, 4) | 3 (3, 4) | 0.427 |
| OGIC | - | - | - | 4 (2, 4) | 3 (3, 4) | 3 (2, 4) | 0.936 |

Median (Q1, Q3)
<sup>1</sup>Chi-squared test used for the number of participants showing $\geq 30\%$ reduction in VAS at the end of the treatment from baseline compared with the average VAS pain intensity during the 7 days prior to study initiation.
Kruskal-Wallis test used for PGIC and OGIC. Global p-values obtained via baseline-adjusted ordinal logistic models for all the other variables.

#### Secondary outcomes

Using a ≥50% reduction in pain VAS scores at the end of the treatment, twelve patients in the Low-Connectivity group (41%), eleven in the High-Connectivity group (37%), and ten in the Classic-M1 group (33%) were responders. Across all secondary outcomes, no between-group differences reached significance (Table 2 and Supplementary Fig. 1). For VAS pain intensity over the last 24 hours, mean pain reduction was 29±49% in the Low-Connectivity group, 32 ± 34% in the High-Connectivity group, and 27±5% in the Classic-M1 group, with no significant group differences (p=0.188). A similar pattern was observed for VAS pain intensity over the last 7 days, with reductions of 18±53%, 13±30%, and 13±28%, respectively (p=0.346). No significant between-group differences were observed for pain severity (p=0.581), pain interference (p=0.954), sleep quality (p=0.180), fatigue (p=0.724), or mood symptoms (p=0.286). Health-related quality of life did not differ significantly across the three treatment arms (p=0.259). PGIC ratings were comparable across groups (p=0.427), as were OGIC ratings (p=0.936).

### Exploratory analyses

Pain VAS reduction after therapy over the last 7 days was associated with local connectivity in the Classic-M1 group (r=0.527, p=0.003; Figure 4A), but not in the Low-Connectivity (r=0.037, p=0.847) or High-Connectivity groups (r=−0.065, p=0.743). Similarly, pain VAS reduction after therapy over the last 24 hours was associated with local connectivity in the Classic-M1 group (r=0.495, p=0.005; Figure 4B), but not in the Low-Connectivity (r=0.279, p=0.136) or High-Connectivity groups (r=−0.205, p=0.296). Interaction analyses of the correlation slope revealed a difference between the Classic-M1 and High-Connectivity groups in regression slopes between local connectivity and pain VAS reduction over the last 24 hours after therapy (p=0.037; Supplementary Table 8).

**Figure 4:**
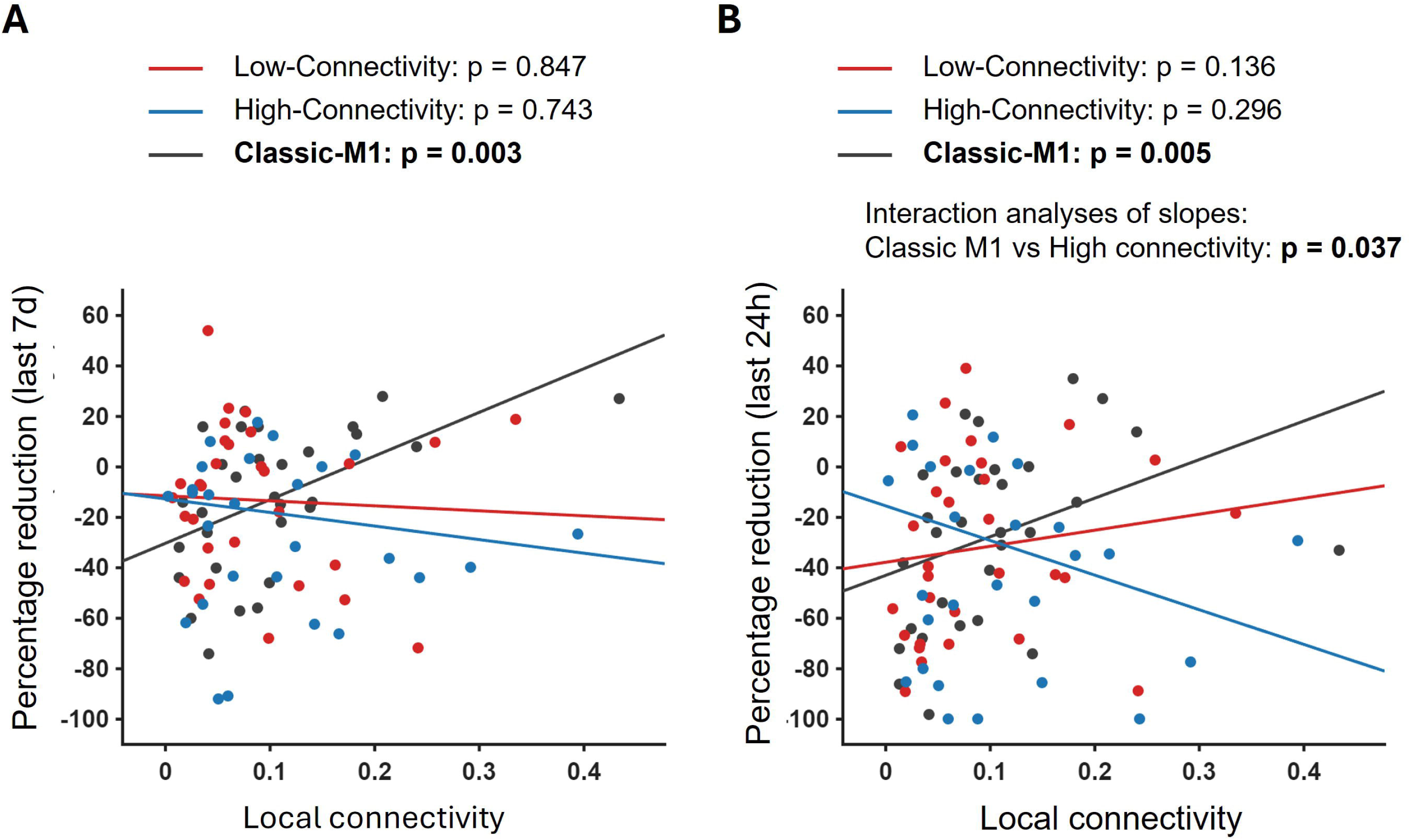
Correlation between local connectivity and percentage pain reduction. **(A)**Correlation between local (within-cluster) connectivity and percentage change in VAS pain intensity over the last 7 days (negative values indicate pain reduction). **(B)** Correlation between local (within-cluster) connectivity and VAS pain reduction in the last 24 hours (negative values indicate pain reduction).

#### Correlation between connectivity index and cortical excitability

The values of the connectivity index and LMFP are reported in Supplementary Table 9. No significant correlation between the connectivity index and LMFP was found (ρ=0.12, p=0.25; Supplementary Fig. 2).

## Discussion

The primary aim of this randomized, double-blind, parallel trial was to determine whether rTMS target allocation based on pre-therapy lower cortical connectivity could identify one of four potential cortical targets associated with a higher probability of response in chronic pain. The results did not show a higher proportion of responders with this strategy compared to classic uninformed M1-rTMS therapy, as no significant differences in pain improvement were observed among the Low-Connectivity, High-Connectivity, and Classic-M1 groups. These findings indicate that a global connectivity index measured after stimulation of four discrete cortical sites was insufficient to capture target-specific neurophysiological markers of rTMS responses in chronic pain. Secondary clinical outcomes also did not differ significantly across the three groups, providing no evidence of a superior analgesic effect for any target.

While global responses elicited by local cortical probing did not inform target selection, exploratory analyses showed that local oscillatory responses following M1 probing did carry predictive value. Specifically, lower pre-therapy local connectivity, measured in the EEG cluster around the stimulation site, was associated with greater pain relief only in the Classic-M1 group. This suggests that local connectivity features of M1 may be more relevant for predicting responsiveness to rTMS than global connectivity measures. These findings open two key research questions in the development of neuromodulation-based brain biomarkers for pain relief: (i) are cortical connectivity biomarkers of rTMS response inherently site-specific? (ii) do local connectivity measures offer greater predictive value than global connectivity indices?

In the present trial, the proportion of patients showing a clinically meaningful reduction in pain intensity was 37% in the Classic-M1 group (11/30 patients), 50% in the High Connectivity group (15/30 patients), and 48% in the Low Connectivity group (14/29 patients). These response rates are consistent with those reported in previous rTMS clinical trials, suggesting that the overall therapeutic efficacy observed in the current study aligns with the existing literature and that connectivity-based target allocation did not enhance analgesic efficacy. Hodaj et al. reported a response rate of 47% (27/57 patients),^12^ Quesada et al. reported a response rate of 47% (17/36 patients)^10^ after M1 rTMS using 30% pain reduction as the definition of responder. Using a 50% of pain reduction threshold, Attal et al. reported a response rate of 44% (22/49 patients).^7^ Investigations of other cortical targets have reported response rates within a comparable range. For PSI stimulation, Dongyang et al. reported clinically meaningful pain reduction in 58% of patients with peripheral neuropathic pain,^22^, whereas Galhardoni et al. observed no clear analgesic effects in patients with central neuropathic pain.^20^ Regarding ACC stimulation, Tzabazis et al. reported a mean pain reduction of 43% in fibromyalgia patients following rTMS,^19^ whereas a larger study in central neuropathic pain failed to demonstrate an analgesic effect.^20^ Results from DLPFC stimulation have also been heterogeneous, with some studies reporting analgesic benefits^16,51,60–62^ and others showing no therapeutic effect.^7^ Therefore, the responder rates of 35–50% and an average pain reduction of approximately 30±40% observed in the current study underscore the persistent challenge of identifying reliable predictors of treatment response, consistent with findings from all previous studies.

A relevant clinical finding of this study concerns the response rate across different chronic pain etiologies. In the present trial, we included patients with different chronic pain conditions, represented by the three mechanistic descriptors of pain: nociceptive, nociplastic, and neuropathic. Yet the observed response rates were comparable to those reported in previous rTMS trials focusing on specific pain conditions such as neuropathic pain and fibromyalgia.^9,10^ This finding suggests that the analgesic effects of rTMS may generalize across chronic pain conditions, rather than being limited to defined mechanistic categories. It also suggests that interindividual differences in cortical connectivity may be more important in predicting response to rTMS than the etiology of pain or its mechanistic descriptors. This is, in itself, not different from other non-pharmacological treatments for chronic pain, which have shown trans-etiological positive effects.^63^

Using TME-EEG, experimental pain models^26^ and chronic pain conditions^64^ have been associated with reductions in cortical excitability and connectivity compared with non-painful conditions or healthy individuals, reflecting altered oscillatory synchronization within pain-related networks, as observed in many resting-state EEG studies in chronic pain.^65^ By probing different cortical regions with TMS, such as M1 and DLPFC, reductions in cortical excitability and connectivity differ across individuals, ^23^ and these differences correlate with individual pain sensitivity.^26,66^ In contrast, rTMS to M1 and PSI have been shown to increase local excitability and connectivity,^27,67^ suggesting that rTMS-induced modulation of cortical local connectivity may be therapeutically relevant, particularly in cortical regions characterized by reduced connectivity.^67^ Based on these results, we hypothesized that cortical targets with the lowest global connectivity would be more susceptible to rTMS-induced analgesia, assuming connectivity–clinical response relationships would be similar across all cortical regions. This hypothesis was not confirmed. Nevertheless, important mechanistic insights emerged from the exploratory analyses. In the Classic-M1 group, pain relief was associated with low local connectivity. This finding suggests that local network organization within M1 may play an important role in determining susceptibility to rTMS-induced modulation. Specifically, lower local phase-based synchronization within M1 may reflect a cortical state with greater capacity for plastic change, thereby facilitating analgesic responses to rTMS. Here, local connectivity was quantified using the debiased wPLI, a phase-based measure that captures the degree to which neuronal populations oscillate with consistent non-zero phase relationships, thereby indexing functional synchronization while minimizing the influence of volume conduction^48^. Lower local connectivity values indicate reduced local phase synchronization, reflecting a more desynchronized cortical network state. In a secondary analysis of the present dataset, comparing responders and non-responders across M1 stimulation, irrespective of arm allocation, and using complementary neurophysiological measures such as ITC and event-related spectral perturbation in the alpha frequency band, we observed that responders to M1 stimulation were characterized by lower pre-therapy local oscillatory synchronization.^68^ Taken together, these findings may indicate that lower pre-therapy local connectivity and oscillatory synchronization within the stimulated M1 region are associated with greater analgesic responses to rTMS, supporting the notion that local M1 network properties may be important to determine M1 rTMS responsivity. Future clinical studies are needed to further investigate and validate this hypothesis.

This study has several notable strengths and limitations. This is the first large, randomized, mechanism-based clinical trial aiming to improve the efficacy of rTMS for chronic pain by using neurophysiological assessment to guide individualized target selection. Whereas most previous studies have focused on comparing active versus sham stimulation or contrasting cortical targets, the present approach aimed to move beyond response-rate estimation toward personalization of treatment based on individual cortical network properties. Second, the exploratory analyses generate testable hypotheses for future trials, suggesting that TMS-EEG-derived local connectivity measures within M1 may help identify patients more likely to respond to rTMS. This study may represent a concrete step toward biomarker-informed patient stratification in non-invasive neuromodulation trials.

Important limitations should also be acknowledged. The study did not include a sham stimulation control group because this was not the trial’s primary objective. The analgesic efficacy of rTMS in chronic pain has been demonstrated in multiple prior sham-controlled studies, and the responder rates observed here are consistent with those reported for active treatment (35-50%). Furthermore, M1 rTMS is currently recommended by international guidelines,^69^ and the use of sham can be ethically questioned. A further limitation concerns whether differences in connectivity values across stimulation targets were driven by variations in local cortical excitability. However, LMFP was not significantly associated with the connectivity index, suggesting that local excitability did not substantially confound this measure. In addition, global connectivity may inherently differ across stimulation targets, potentially influencing the distribution of DLPFC and PSI targets between connectivity groups. This highlights the potential value of more localized connectivity measures to mitigate such bias.

In conclusion, this study showed that rTMS target allocation based on a global cortical connectivity index derived from TMS–evoked EEG potentials did not significantly identify a cortical target associated with a higher probability of treatment response. Exploratory analyses revealed that lower pre-therapy local connectivity within the stimulated M1 region was associated with greater pain relief. Future studies should prospectively test M1-local connectivity–based stratification to better optimize individualized neuromodulation strategies for chronic pain.

## Acknowledgements

The current study was supported by an ERC Horizon Europe Consolidator grant (PersoNINpain 101087925) and a Novo Nordisk grant (Grant NNF21OC0072828). The Center for Neuroplasticity and Pain was supported by the Danish National Research Foundation (DNRF121). TGN receives funding from the Lundbeck Foundation (R441-2023-232).

## Author contribution

**Enrico De Martino:** Conceptualization, Methodology, Formal analysis, Investigation, Data curation, Visualization, Writing - Original draft preparation; Project administration; **Margit Midtgaard Bach**: Investigation, Data curation, Writing - Review & Editing; **Anne Jakobsen**: Investigation, Writing - Review & Editing; **Bruno Andry Nascimento Couto**: Software, Formal analysis, Data Curation, Visualization, Writing - Review & Editing; **Pedro Nascimento Martins**: Visualization, Statistical analysis, Writing - Review & Editing; **Adenauer Girardi Casali**: Software; Writing - Review & Editing; **Stian Ingemann-Molden**: Investigation, Writing - Review & Editing; **Thomas Graven-Nielsen**: Conceptualization, Methodology, Writing - Review & Editing; **Daniel Ciampi de Andrade**: Conceptualization, Methodology, Visualization, Writing - Review & Editing; Project administration, Funding acquisition.

## Data availability

The data of this study are available from the corresponding author upon reasonable request.

