## Supplemental material for "Personalizing neuromodulation for chronic pain: A connectivity-guided randomized trial"

### Supplementary material

#### *Supplementary Table 1*

The clusters of EEG electrodes used for the exploratory analysis.

| Target | Side | Electrodes |
|---|---|---|
| M1 | Left | C1, C3, C5, Cp1, Cp3, Cp5 |
| | Right | C2, C4, C6, Cp2, Cp4, Cp6 |
| DLPFC | Left | F1, F3, F5, Fc1, Fc3, Fc5 |
| ACC | Left | FCz, Cz, FC1, FC2, C1, C2 |
| | Right | FCz, Cz, FC1, FC2, C1, C2 |
| PSI | Left | FC7, FC3, C7, C5, C3 |
| | Right | FC8, FC4, C8, C6, C4 |

#### *Supplementary Table 2*

Main diagnosis of 90 chronic pain patients.

| **Classic M1**<br>N = 30 | **High Connectivity**<br>N = 30 | **Low Connectivity**<br>N = 30 |
|---|---|---|
| Post-traumatic foot CRPS type 2 with neuropathy | Shoulder arthropathy | Traumatic radiculopathy |
| Traumatic nerve lesion (ulnar nerve) | Spinal cord injury | Hereditary erythromelalgia |
| Post traumatic spondylopathy | Traumatic radiculopathy | Diabetic polyneuropathy |
| Chronic widespread pain | Postoperative neuropathy | Knee osteoarthrosis |
| Cervical traumatic radiculopathy | Cervical spondylopathy | Pudendal neuropathy |
| Knee osteoarthrosis | CRPS I | Traumatic lumbar radiculopathy |
| Fibromyalgia | Shoulder arthropathy | Chronic primary abdominal pain syndrome |
| Neck arthrosis | Chronic primary abdominal pain syndrome | Fibromyalgia |
| Fibromyalgia | Fibromyalgia | Erythromelalgia |
| Fibromyalgia | Lumbar spine degenerative spondyloarthropathy | Fibromyalgia |
| Fibromyalgia | Fibromyalgia | Chronic primary myofascial temporomandibular disorder pain |
| Inflammatory bowel disease-related arthropathy | Polyneuropathy | Postoperative arthropathy |
| Diabetic polyneuropathy | Radiculopathy | Fibromyalgia |
| Fibromyalgia | Osteoarthritis | Fibromyalgia |
| Diabetic polyneuropathy | Diabetic polyneuropathy | osteoarthritis |
| Shoulder pain with arthropathy | Fibromyalgia | Chronic primary temporomandibular disorder pains |
| Fibromyalgia | Polyneuropathy | Fibromyalgia |
| Fibromyalgia | Chronic widespread pain | Chronic primary cervical pain |
| Chronic primary cervical pain | Fibromyalgia | Post-operative arthropathy |
| Fibromyalgia | Fibromyalgia | Fibromyalgia |
| Fibromyalgia | post traumatic | Systemic Lupus Erythematosus with arthritis |
| Arthropathy | Post traumatic spondyloarthropathy | Lumbar spine degenerative spondyloarthropathy |
| Post-operative | Cervical spine degenerative spondyloarthropathy | Chronic primary cervical pain |
| Osteoarthritis | Brachial plexus avulsion | Polyneuropathy |
| Chronic widespread pain | Lumbar spine degenerative spondyloarthropathy | Ulnar neuropathy |
| Polyneuropathy | Fibromyalgia | Arthritis due to Sjogren's disease |
| Knee osteoarthrosis | Fibromyalgia | Polyneuropathy |
| Lumbar spinal stenosis with radiculopathy | Peripheral nerve lesion | Hip arthropathy |
| Lumbar spinal stenosis with radiculopathy | Inflammatory knee arthritis | Traumatic radiculopathy |

#### *Supplementary Table 3*

Medications used before and after treatment.

| Medication | Group | Before rTMS | After rTMS |
|---|---|---|---|
| Acetaminophen | Low-Connectivity | 18 | 15 |
| | High-Connectivity | 15 | 13 |
| | Classic-M1 | 23 | 16 |
| NSAIDs | Low-Connectivity | 12 | 8 |
| | High-Connectivity | 10 | 9 |
| | Classic-M1 | 11 | 10 |
| Baclofen | Low-Connectivity | 1 | 1 |
| | High-Connectivity | 0 | 1 |
| | Classic-M1 | 1 | 0 |
| Pregabalin | Low-Connectivity | 3 | 4 |
| | High-Connectivity | 4 | 4 |
| | Classic-M1 | 5 | 5 |
| Gabapentin | Low-Connectivity | 3 | 2 |
| | High-Connectivity | 2 | 4 |
| | Classic-M1 | 6 | 6 |
| Morphin | Low-Connectivity | 3 | 3 |
| | High-Connectivity | 1 | 1 |
| | Classic-M1 | 3 | 4 |
| Tramadol | Low-Connectivity | 1 | 0 |
| | High-Connectivity | 3 | 1 |
| | Classic-M1 | 7 | 4 |
| Codeine | Low-Connectivity | 0 | 0 |
| | High-Connectivity | 2 | 0 |
| | Classic-M1 | 0 | 0 |
| Naloxone | Low-Connectivity | 2 | 3 |
| | High-Connectivity | 2 | 2 |
| | Classic-M1 | 2 | 2 |
| Naltrexone | Low-Connectivity | 5 | 6 |
| | High-Connectivity | 5 | 5 |
| | Classic-M1 | 4 | 4 |
| Amitriptyline | Low-Connectivity | 4 | 3 |
| | High-Connectivity | 2 | 2 |
| | Classic-M1 | 0 | 0 |
| Nortriptyline | Low-Connectivity | 1 | 1 |
| | High-Connectivity | 2 | 1 |
| | Classic-M1 | 0 | 0 |
| Duloxetine | Low-Connectivity | 2 | 4 |
| | High-Connectivity | 1 | 1 |
| | Classic-M1 | 3 | 3 |
| Venlafaxine | Low-Connectivity | 0 | 0 |
| | High-Connectivity | 1 | 1 |
| | Classic-M1 | 0 | 0 |

| | | | |
|---|---|---|---|
| Citalopram | Low-Connectivity | 0 | 0 |
| | High-Connectivity | 1 | 1 |
| | Classic-M1 | 0 | 0 |
| Mirtazapine | Low-Connectivity | 0 | 0 |
| | High-Connectivity | 0 | 0 |
| | Classic-M1 | 1 | 0 |

#### *Supplementary Table 4*

Stimulation intensities used for TMS–EEG and rTMS presented as mean ± standard deviation.

| Group | TMS- EEG | rTMS intensity first session | rTMS intensity last session |
|---|---|---|---|
| Low-Connectivity | 57±10% | 46±9% | 47±6% |
| High-Connectivity | 55±12% | 45±10% | 45±8% |
| Classic-M1 | 50±6% | 41±6% | 41±6% |

### *Supplementary Table 5*

Side effects during the rTMS treatments

| Side effect | Classic M1 | High connectivity | Low connectivity | p-value[1] |
|---|---|---|---|---|
| Unusual smells, tastes, or emotions | 0 (0%) | 2 (6.7%) | 3 (10%) | 0.363 |
| Unusual experiences – 'out of body experiences', feeling distracted, body feels different | 2 (6.7%) | 0 (0%) | 1 (3.3%) | 0.770 |
| Dizziness | 4 (13%) | 8 (27%) | 11 (37%) | 0.115 |
| Twitching movements of an arm, leg, or other body part | 1 (3.3%) | 4 (13%) | 6 (20%) | 0.171 |
| Trembling, numb sensation, or feeling of electricity in a part of the body | 3 (10%) | 4 (13%) | 3 (10%) | >0.999 |
| Headache | 9 (30%) | 12 (40%) | 11 (37%) | 0.712 |
| Unexplained confusion, drowsiness, or weakness | 1 (3.3%) | 0 (0%) | 2 (6.7%) | 0.770 |

[1]Fisher's exact test; Pearson's Chi-squared test

#### *Supplementary Table 6*

Likelihood-ratio tests for mixed-effects models

| Predictor | Chi-Square | P |
|---|---|---|
| **Connectivity index** | | |
| Target connectivity | 0.09 | 0.958 |
| Target connectivity (nonlinear) | 0.08 | 0.775 |
| Baseline pain | 7.82 | 0.020 |
| Baseline pain (nonlinear) | 0.72 | 0.398 |
| Model (nonlinear terms) | 0.76 | 0.683 |
| Model (total) | 7.97 | 0.093 |
| **Local-to-M1 difference (LTM1D)** | | |
| LTM1D (target–M1) | 0.60 | 0.740 |
| LTM1D (nonlinear) | 0.46 | 0.499 |
| Baseline pain | 8.03 | 0.018 |
| Baseline pain (nonlinear) | 0.82 | 0.364 |
| Model (nonlinear terms) | 1.22 | 0.544 |
| Model (total) | 8.47 | 0.076 |

#### *Supplementary Table 7*

Global connectivity index in the Low-Connectivity, High-Connectivity, and Classic-M1 groups.

| Group | Global connectivity index |
|---|---|
| Low-Connectivity | 0.81 ± 0.57 |
| High-Connectivity | 3.81 ± 3.79 |
| Classic-M1 | 1.83 ± 2.54 |

#### *Supplementary Table 8*

Interactions of local connectivity with percentage pain visual analogue scale (VAS) reduction in the last 7 days and the last 24 hours.

| Analysis | Contrast | Interaction (95% CI) | p-value |
|---|---|---|---|
| Local connectivity and pain VAS in the last 7 days | Classic-M1 – High-Connectivity | 220 (-7, 448) | 0.058 |
| | Classic-M1 – Low-Connectivity | 193 (-49, 434) | 0.118 |
| | High-Connectivity – Low-Connectivity | -27 (-264, 210) | 0.821 |
| Local connectivity and pain VAS in the last 24 hours | **Classic-M1 – High-Connectivity** | **280 (16, 543)** | **0.037** |
| | Classic-M1 – Low-Connectivity | 89 (-190, 369) | 0.531 |
| | High-Connectivity – Low-Connectivity | -190 (-465, 84) | 0.174 |

#### *Supplementary Table 9*

Connectivity index and local mean-field power for each target (n = 90).

| | Local connectivity index | Local mean field power |
|---|---|---|
| M1 | 1.83 ± 2.65 | 0.19 ± 0.09 |
| DLPFC | 1.06 ± 1.03 | 0.12 ± 0.06 |
| ACC | 1.91 ± 2.07 | 0.24 ± 0.14 |
| PSI | 2.09 ± 2.08 | 0.18 ± 0.11 |

#### *Supplementary Figure 1*

Secondary clinical measures assessed before and after rTMS treatment. Bar plots display group means ± SD with individual data points overlaid. **(A)** Pain intensity in the last 24 h, **(B)** pain intensity in the last week, **(C)** BPI pain severity, **(D)** BPI pain interference, **(E)** number of body areas with pain, **(F)** number of medications, **(G)** sleep quality, **(H)** fatigue, **(I)** Hospital Anxiety and Depression Scale (HADS), and **(J)** EQ-5D quality-of-life score.

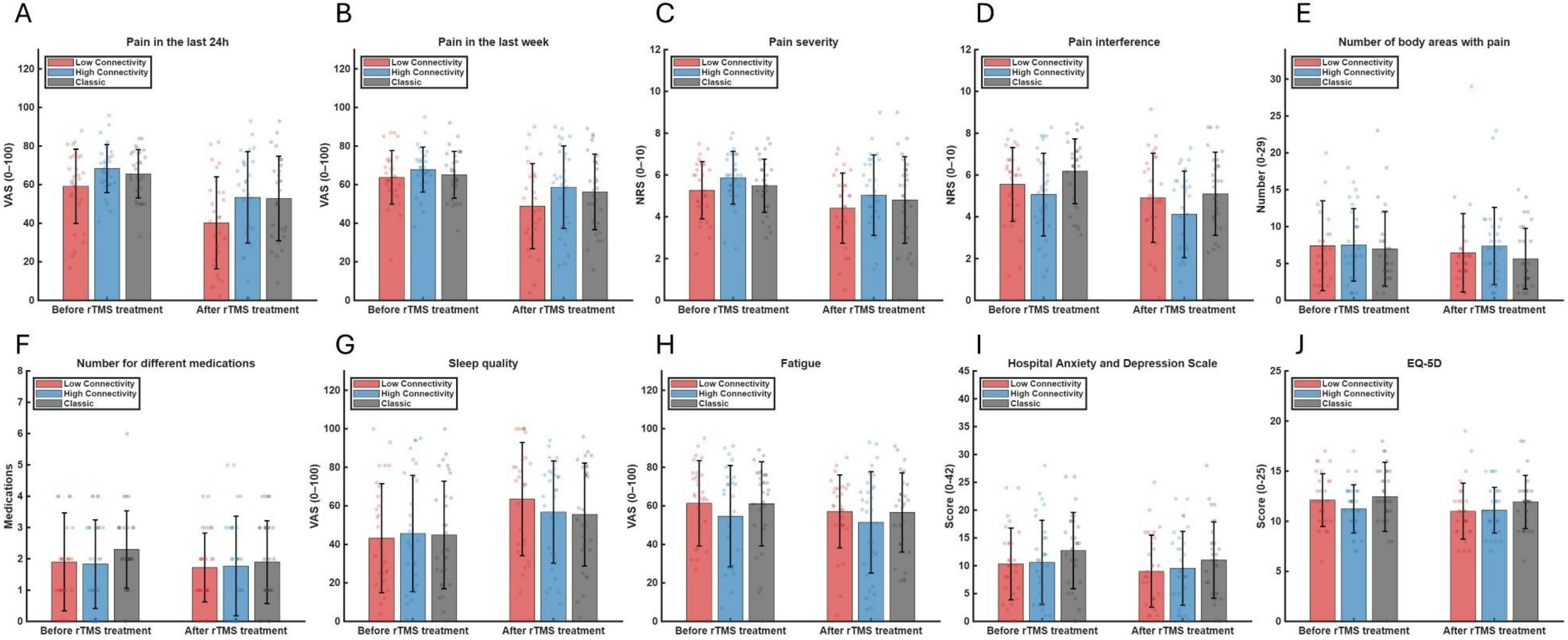

#### *Supplementary Figure 2*

Correlation between the local connectivity index and local mean field power.

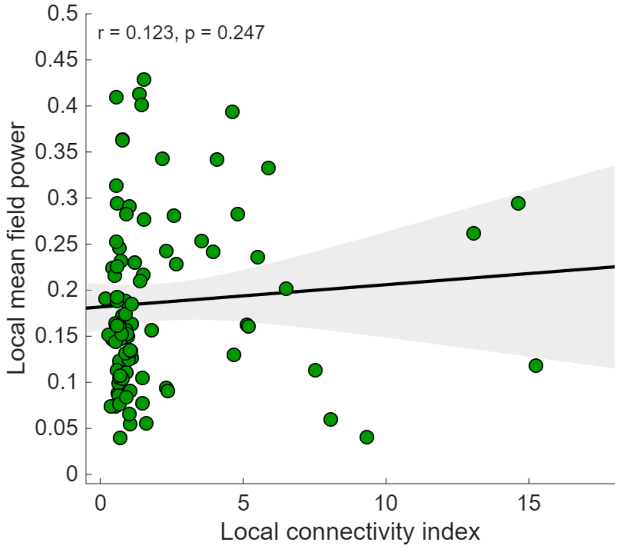
